# Single Cell Transcriptional Archetypes of Airway Inflammation in Cystic Fibrosis

**DOI:** 10.1101/2020.03.06.20032292

**Authors:** Jonas C. Schupp, Sara Khanal, Jose L. Gomez, Maor Sauler, Taylor S. Adams, Geoffrey L. Chupp, Xiting Yan, Sergio Poli, Ruth R. Montgomery, Ivan O. Rosas, Charles S. Dela Cruz, Emanuela M. Bruscia, Marie E. Egan, Naftali Kaminski, Clemente J. Britto

## Abstract

Cystic fibrosis (CF) is a life-shortening multisystem hereditary disease caused by abnormal chloride transport. CF lung disease is driven by innate immune dysfunction that perpetuates inflammation. The airways provide a window into CF pathogenesis, as immune cells display exaggerated inflammatory responses and impaired phagocytic function, contributing to tissue injury. In order to define the transcriptional profile of this airway immune dysfunction, we performed the first single-cell transcriptome characterization of CF sputum. We show that the airway immune cell repertoire shifted from alveolar macrophages in HC to a predominance of recruited monocytes and neutrophils in CF. Recruited lung mononuclear phagocytes were abundant in CF, separated into three archetypes: activated monocytes, monocyte-derived macrophages, and heat-shock activated monocytes. Neutrophils were most prevalent in CF, with a dominant immature proinflammatory archetype. While CF monocytes exhibited proinflammatory features, both monocytes and neutrophils showed transcriptional evidence of abnormal phagocytic and cell-survival programs. Our findings offer an opportunity to understand subject-specific immune dysfunction and its contribution to divergent clinical courses in CF. As we progress towards personalized applications of therapeutic and genomic developments, we hope this inflammation profiling approach will enable further discoveries that change the natural history of CF lung disease.

## Introduction

Cystic Fibrosis (CF) is a life-shortening, multi-organ autosomal recessive disease that affects approximately 75,000 patients worldwide. In the United States, CF is the most common fatal genetic disease, affecting over 33,000 patients (*1, 2*). Clinical manifestations of CF are caused by a mutation in the cystic fibrosis transmembrane conductance regulator (CFTR) gene that causes abnormal chloride and bicarbonate transport on epithelial surfaces of the respiratory and gastrointestinal tracts (*3*). The disruption of epithelial and innate immune functions are key contributors to the development of CF lung disease (CFLD), the primary cause of morbidity and mortality in CF (*4-6*). Immune dysfunction in CF extends beyond *CFTR* expression to include numerous disease-modifying genes that contribute to its clinical phenotype and progression (*7, 8*).

Chronic airway inflammation is crucial in the development of CFLD, where recruited inflammatory cells cause tissue damage and contribute to airway remodeling (*9, 10*). These inflammatory cell populations are heterogenous, with CF-specific polymorphonuclear neutrophil (PMN) and macrophage (MΦ) subclasses being increasingly recognized (*10, 11*). Previous studies of inflammatory cells in CF have profiled immune cells from blood and lung biopsies using bulk RNA sequencing to characterize gene expression profiles associated with disease progression and clinical outcomes (*12*). Flow-cytometry-based studies have also contributed greatly to our understanding of functional defects in subsets of CF inflammatory cells (*10, 13*). Yet, a single-cell transcriptome characterization of CF airway cells has not been reported.

Sputum is an ideal sample to understand host-pathogen and cell-cell interactions within the airway. In CF, respiratory secretions provide non-invasive access to the primary compartment at the center of CFLD pathogenesis. Sputum cells reflect the complex airway interactions between inflammatory cells, pathogens, and the CF airway microenvironment. These interactions can only be partially investigated in ex vivo human models, sorted cell populations, or in biopsies of terminally diseased tissues.

Airway neutrophilic inflammation in CF has been well characterized in sputum (*14, 15*). CF PMN have a proinflammatory profile, yet some studies reveal functionally different subsets of PMNs, including populations with abnormal immune function and defective bacterial killing (*10, 11*). Airway MΦ and other mononuclear phagocytes are also present in human CF airway secretions (*16, 17*). As a group, airway monocytes (Mo) in CF have impaired phagocytic function and enhanced cytokine production, however their role in CF pathogenesis is not fully understood (*18-20*). Mo appear to play an important role in driving exaggerated airway inflammation in animal models of CF (*4, 9, 16, 19*), however a more granular approach to characterize mononuclear phagocyte populations according to states of maturity and activation has not been accomplished in sputum.

Single-cell transcriptome profiling is a powerful tool to study innate immune defects and define key cell subpopulations that contribute to CFLD pathogenesis. The prior identification of discrete inflammatory cell subpopulations in CF suggested to us that these cells do not exist in separate clusters in the airways, but rather as a continuum of immune maturation and function. To define the spectrum of maturation and immune activation of inflammatory cells CF sputum, we applied single-cell RNA sequencing (scRNAseq) followed by Potential of Heat diffusion for Affinity-based Transition Embedding (PHATE) and Pseudotime analysis, novel approaches to visualize high-dimensional data. In the continuum of sputum inflammatory cells, those with most extreme gene expression features defined functional and maturity trajectories, here called transcriptional archetypes (*21*). These archetypes constitute a dynamic, more inclusive way to understand gene expression differences between airway cells.

This work is the first to fully characterize the spectrum of maturation and immune activation states of inflammatory cell populations in CF airways at an unprecedented genomic level of resolution enabled by scRNAseq. Our study underscores the advantages of high-throughput approaches to characterize cell populations in a diseased tissue or compartment, or in identifying differential functional activation patterns. The development of transcriptional inflammatory cell archetypes could identify novel cell types and gene expression differences responsible for divergent clinical courses in subjects with similar CF-causing mutations but different airway inflammatory profiles, opening the door for highly-targeted therapeutic interventions.

## Results

### Disease-Specific Cell Distributions of CF Airway Inflammatory and Epithelial Cells

The primary objective of this study was to characterize sputum cell subpopulations in CF using unbiased transcriptome analysis of single cells obtained CF and healthy control (HC) subjects. Our recruitment period extended from December 2018 through December 2019. Nine subjects with a confirmed CF diagnosis from the Yale Adult CF Program provided sputum samples. We also recruited five HC to undergo sputum induction according to previous protocols (*22*).

Study subjects were closely age-matched, with a higher inclusion of female subjects in the CF group (67% CF, n=6, 40% HC, n=2). The CF cohort was comprised primarily of *F508del* homozygous subjects (78%, n=7) with only two *F508del* heterozygotes harboring either one deletion or one frameshift mutation in one *CFTR* allele and an *F508del* in the other. The CF cohort’s degree of lung function impairment, as determined by Forced Expiratory Volume in the first second (FEV_1_), ranged from mild to severe (FEV_1_ 19-84% of predicted), with a mean FEV_1_ of 57%. All CF subjects had pancreatic exocrine insufficiency and 44% (n=4) carried a diagnosis of CF-related diabetes. *Pseudomonas aeruginosa* was isolated in the sputum of 56% of CF subjects (n=5). The majority of CF subjects were receiving CFTR-modulator therapy (89%, n=8) with a combination of either Ivacaftor/Tezacaftor (67%, n=6) or Ivacaftor/Lumacaftor (22%, n=2). For further demographic and clinical details see Table 1.

**Table 1.** Demographic characteristics of study subjects from the Yale Adult Cystic Fibrosis Program and healthy controls. HC: Healthy controls; CF: CF subjects; FEV_1_ Forced expiratory volume in the first second; BMI: Body Mass Index; PI: Pancreatic Exocrine Insufficiency; CFRD: CF-related Diabetes; PA: Pseudomonas aeruginosa; CFTR: Cystic Fibrosis Transmembrane conductance Regulator.

| *Number of Patients (n)* | *HC (5)* | *CF (9)* |
| --- | --- | --- |
| Age |  |  |
| Age (Mean) | 35.4 | 30.6 |
| Age (STD) | 5.9 | 6.5 |
| Age (Range) | 26-42 | 24-43 |
| Sex |  |  |
| Female (n) | 2 | 6 |
| Female (%) | 40 | 67 |
| Male (n) | 3 | 3 |
| Male (%) | 60 | 33 |
| Mutation Background |  |  |
| F508del/F508del (n) | NA | 7 |
| F508del/F508del (%) | NA | 77.8 |
| F508del/other (n) | NA | 2 |
| F508del/other (%) | NA | 22.2 |
| No *F508del* mutations (n) | NA | 0 |
| No *F508del* mutations (%) | NA | 0 |
| FEV<sub>1</sub> (L) |  |  |
| FEV<sub>1</sub> (Mean) | NA | 1.9 |
| FEV<sub>1</sub> (STD) | NA | 0.7 |
| FEV<sub>1</sub> (Range) | NA | 0.68 - 2.85 |
| FEV<sub>1</sub> (%) |  |  |
| FEV<sub>1</sub> (Mean) | NA | 57 |
| FEV<sub>1</sub> (STD) | NA | 21.5 |
| FEV<sub>1</sub> (Range) | NA | 19 - 84 |
| BMI (Kg/m<sup>2</sup>) |  |  |
| BMI (Mean) | NA | 22.2 |
| BMI (STD) | NA | 2.1 |
| BMI (Range) | NA | 19.11 - 25.73 |
| CF Comorbidities |  |  |
| PI (n) | NA | 9 |
| PI (%) | NA | 100 |
| CFRD (n) | NA | 4 |
| CFRD (%) | NA | 44.4 |
| Liver disease (n) | NA | 1 |
| Liver disease (%) | NA | 11.1 |
| Microbiology |  |  |
| PA Colonization (n) | NA | 5 |
| PA Colonization (%) | NA | 55.6 |
| CFTR Modulators |  |  |
| Ivacaftor/Tezacaftor (n) | NA | 6 |
| Ivacaftor/Tezacaftor (%) | NA | 66.7 |
| Ivacaftor/Lumacaftor (n) | NA | 2 |
| Ivacaftor/Lumacaftor (%) | NA | 22.2 |
| No modulator (n) | NA | 1 |
| No modulator (%) | NA | 11.1 |

We developed a standardized scRNAseq workflow for sputum sample analysis (Fig. 1A) and profiled a total of 20,095 sputum cells (12,494 CF, 7,601 HC). We identified nine distinct sputum cell populations based on known genetic markers (Fig. 1C, Data file S1): mononuclear phagocytes (recruited lung Mo, Mo-derived MΦ (MoMΦ), and alveolar MΦ (alvMΦ)); classical and plasmacytoid dendritic cells (cDC, pDC); PMN; lymphocytes (B, T, and NK cells); and airway epithelial cells from buccal and tracheobronchial mucosa (Fig. 1B-D).

**Fig.1.**
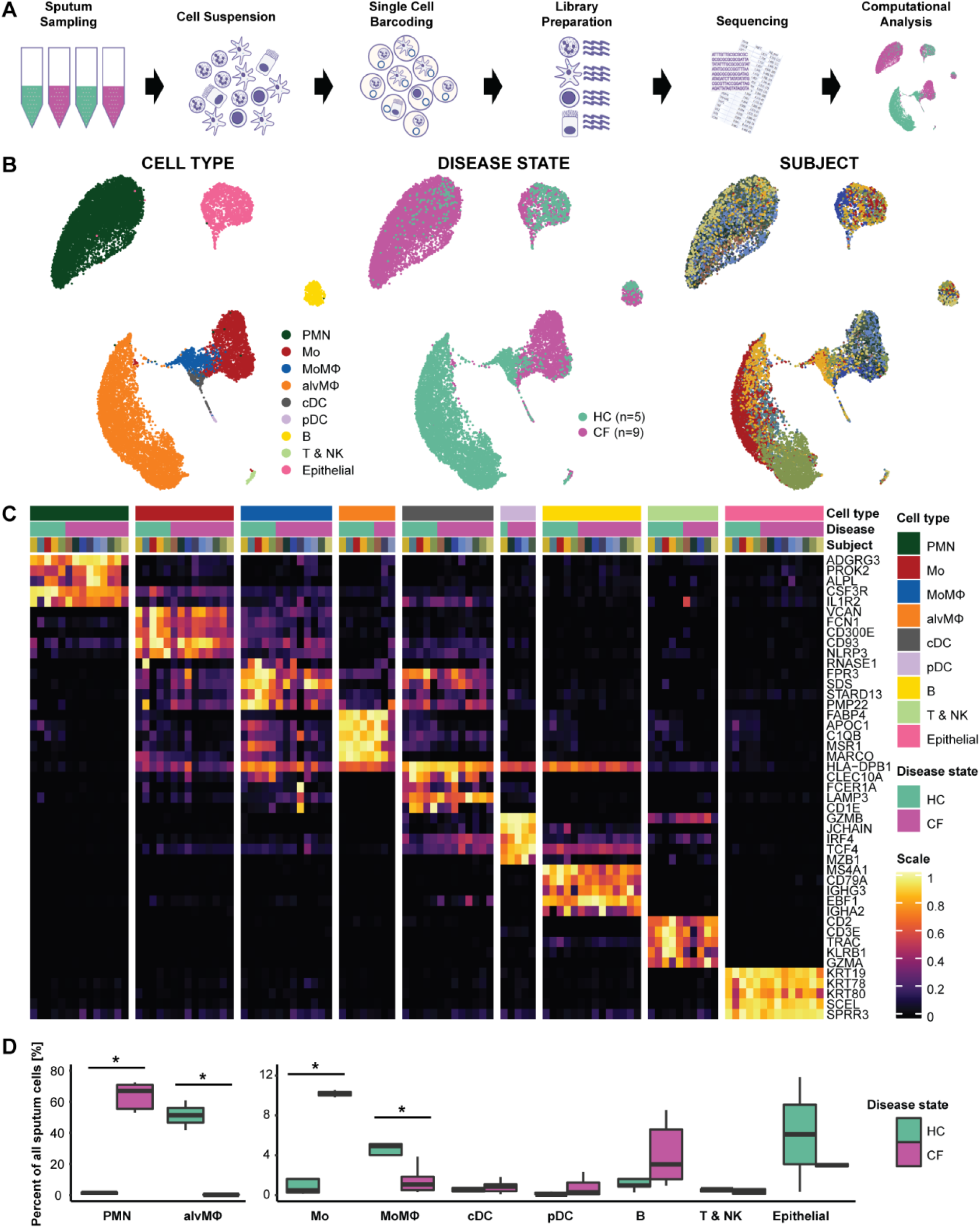
ScRNAseq Reveals an Immune Cell Repertoire Shift from Alveolar MΦ to Recruited Monocytes and PMN in CF. **(A)** Schematic of the experimental design. (i) Spontaneously expectorated sputum from patients with cystic fibrosis (CF) and induced sputum from healthy controls (HC) was collected. (ii) Sputum was processed into a single-cell suspension. (iii) Droplet-based scRNAseq barcoding (iii) library preparation (iv) sequencing (v) and computational analysis. **(B)** Uniform Manifold Approximation and Projection (UMAP) visualization of 20,095 sputum cells from nine patients with CF and five controls. Each dot represents a single cell, and cells are labelled by (i) cell type, (ii) disease status, and (iii) subject. **(C)** Heatmap of marker genes for all cell types identified. Each column represents the average expression value of one subject, grouped by disease status and cell type. Gene expression values are unity-normalized from 0 to 1. **(D)** Boxplots showing percentages of all identified cell types to all cells profiled per subject, separated by disease state. Whiskers represent 1.5 × interquartile range (IQR). * p < 0.05 determined by a Wilcoxon rank sum test comparing cell percentages of CF patients and controls. Mo: Monocyte; MoMΦ: monocyte-derived macrophage; alvMΦ: Alveolar macrophage; cDC: classical dendritic cell, pDC: plasmacytoid dendritic cell; B: B-lymphocyte; T & NK: T-lymphocytes and NK-cells; PMN: polymorphonuclear neutrophil.

### The Inflammatory Cell Repertoire of CF Sputum Displays a Shift from alvMΦ to Airway Monocytes and PMN

The dominant cell populations in CF and HC samples were strikingly different. PMNs contributed 64% of all CF cells, with minimal numbers of alvMΦ (0·4%). In contrast HC samples were composed of 80·2% alvMΦ with almost no detectable PMN (<2%, both p <0·002). Further, CF subjects also exhibited increased numbers of airway Mo (19% CF, 1% HC, p=0·001) and B cells (4% CF, 1% HC, p = ns), and lower numbers of MoMΦ (1% CF, 6% HC, p=0·007) (Fig. 1B-D). Disease-associated PMN, MΦ, and Mo cellular distributions were confirmed on mass cytometry data from a previously published study by our group, comparing surface markers of inflammatory sputum cells in CF and HC (Fig. S1) (*22*). Furthermore, correlation of cell type gene classifiers in this study and analogous cell types in the largest scRNAseq dataset of the distal lung (n=28) revealed a greater correlation between HC cell types from each dataset than within other cell types from the same dataset, confirming our cell annotations (Fig. S2) (*23*). Our findings indicate that immune cell populations in CF sputum are distinguishable from HC through scRNAseq, and that our cell annotations and shifts in major cell distributions in CF are consistent with other mass cytometry (CyTOF) and scRNAseq studies.

### Recruited CF Lung Mononuclear Phagocytes Display Distinct Maturation and Immune Activation Archetypes

AlvMΦ were rare in CF sputum; however, we identified a distinct subpopulation of Recruited Lung mononuclear Phagocytes (RLPs, Fig. 1B) that included recruited lung Mo and MoMΦ. These RLPs were defined by high expression of mononuclear phagocyte-associated genes (*LYZ, CTSB, CTSH, CTSL, CTSS, CTSZ, HLA-DRA, HLA-DRB1, LGALS1, FTL, CD74*). RLPs were relatively abundant in CF (20% of CF cells) and were rarely identified in HC sputum (7% of HC cells, p=0·06). RLPs were a heterogeneous group, with pronounced and notably different plasticity in CF. This suggested that RLPs would differ not only in abundance, but also in transcriptional profiles between HC and CF.

To characterize the spectrum of immune activation and maturation of Mo and MoMΦ contained within CF and HC RLPs, we performed a Pseudotime analysis using PHATE. Pseudotime (trajectory inference) is a computational technique that allows the distribution of single-cell expression profiles along the continuum of a biologic process marked by gene expression changes (in this case cell maturation, immune activation, and heat-shock response gene expression). Pseudotime analysis demonstrated three distinct gene expression trajectories, and in turn, the most extreme phenotypes of these trajectories defined three RLP transcriptional archetypes in sputum (Fig. 2A)(*21*). Two of these archetypes were CF-predominant archetypes: activated proinflammatory Mo and heat-shock activated Mo (HS-Mo). The third RLP archetype, mature resting MoMΦ, was more prevalent in HC.

**Fig.2.**
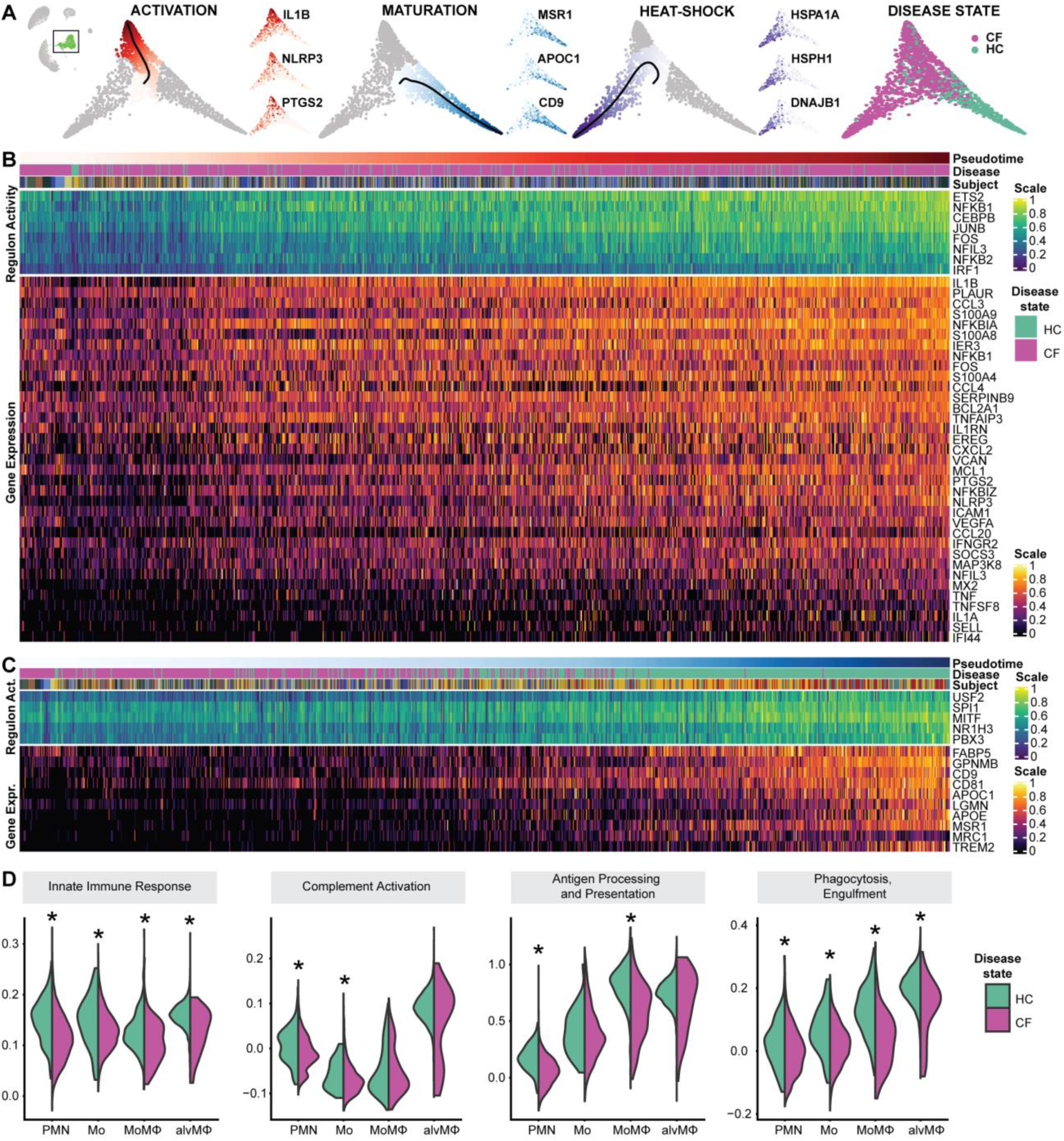
Recruited Lung Mononuclear Phagocytes are a Distinct Cell Population with a Broad Spectrum of Maturity and Immune Activation in CF Airways. **(A)** Potential of Heat diffusion for Affinity-based Transition Embedding (PHATE) of monocytes and monocyte-derived macrophages, colored by pseudotime, all starting from quiescent monocytes towards (i) activated monocytes, (ii) mature monocyte-derived macrophages, (iii) monocytes expressing a heat-shock response. (iv) monocytes and monocyte-derived macrophages, colored by disease state. All three archetypes are accompanied by three PHATE plots colored by the gene expression of typical genes ramping up along a specific pseudotime. For corresponding UMAP embedding colored by gene expressions of the same genes, see Fig. S3. For corresponding PHATE embedding colored by cell type and subjects, see Fig. S4. **(B)** Heatmap of gene expression and regulon activity in monocytes undergoing activation, ordered by pseudotime distances along PHATE manifolds that transition from quiescent monocytes towards an activated monocyte archetype. **(C)** Heatmap of gene expression and regulon activity in monocytes undergoing maturation, ordered by pseudotime distances along PHATE manifolds that transition from quiescent monocytes towards a control-enriched mature monocyte-derived macrophage archetype. In both heatmaps: annotation bars represent the pseudotime distance, disease status, and subject for each cell; expression values are centered and scaled. **(D)** Violin plots of pathway activity scores, grouped by cell type, separated by disease state. Depicted pathway scores from left to right are: GO:0045087 - innate immune response, GO:0006958 - complement activation, classical pathway, GO:0019882 - antigen processing and presentation, GO:0006911 - phagocytosis, engulfment. Mo: Monocyte; MoMΦ: monocyte-derived macrophage; alvMΦ: Alveolar macrophage; PMN: polymorphonuclear neutrophil.

Next, we examined the sequence of gene expression changes leading to the mature resting MoMΦ and activated proinflammatory Mo archetypes, correlating gene expression changes with Pseudotime distance values. The trajectory towards activated proinflammatory Mo was characterized by a gradual and steady increase of proinflammatory chemokine and cytokine gene expression. This trajectory was characterized by increasing expression of *IL1B, CXCL2, CCL3, CCL4, CCL20, VEGFA* and *EREG*, Calprotectin (*S100A8, S100A9*), anti-apoptotic proteins *MCL1* and *BCL2L1*, the inflammasome subunit *NLRP3*, inducible cyclooxygenase 2 (*PTGS2*), and transcription factor *NFKB1* (Fig. 2B, Data file S2). In the activated Mo archetype, imputed regulating factors of common activator/repressor genes (i.e. regulons), suggested increased expression of *NFKB1* and proinflammatory transcription factors *NFKB2, ETS*, and *IRF1*. Proinflammatory cytokines *TNF* and *IL1A* were expressed only towards the extreme end of the trajectory, in the most activated Mo. In contrast to CF RLPs, we did not observe similar immune activation archetypes in MoMΦ, or in alvMΦ from HC. Remarkably, although proinflammatory CF Mo exhibited increased overall cytokine expression, they also showed impaired expression of key phagocytic and cytolytic components of the immune response (complement C1Q), markers of maturation towards a MΦ phenotype (*APOC1, APOE*), and phagocytic function (*MARCO*) compared to other RLP archetypes (Fig. 2B, D).

The mature resting MoMΦ archetype was enriched in HC, and none of the CF MΦ reached the distal end of this archetype (Fig. 2C). Key regulons involved in Mo to MΦ maturation were active, and increasingly expressed towards the distal end of the archetype trajectory, including canonical *SPI1* (*PU*.*1*), as well as *MITF* and *USF2*. Maturation of MoMΦ was accompanied by a gradual transcriptional increase of scavenger and pattern-recognition receptors *MSR1* and *MRC1*, surface markers *CD9* and *CD81*, apolipoproteins *APOC1* and *APOE*, and *FABP5*.

MoMΦ were overall rare in sputum, but more evenly distributed between CF and HC subjects, these were distinguished by expression of *PLA2G7*, an enzyme that inactivates platelet-activating factor, monocyte chemokine *CCL2, LGMN* a cysteine-protease involved in MHC-II presentation and differentiation towards DC and activated-leukocyte cell adhesion molecule *ALCAM*. The majority of sputum cells in HC were alvMΦ. These alvMΦ were indistinguishable from those found in the distal lungs of HC volunteers in a large-scale scRNAseq study of lung cells in idiopathic pulmonary fibrosis and control lungs (Fig. S2)(*23*). These highly abundant HC alvMΦ expressed the expected levels of phagocytosis-associated genes, underscoring the transcriptional readiness of healthy immune cells to participate in phagocytic functions and coordinate inflammatory cell recruitment, without the basal proinflammatory activity noted in the CF-predominant Mo. Taken together, these findings show that CF RLPs have high proinflammatory gene expression but limited phagocytosis-associated transcriptional responses, consistent with excessive inflammation and impaired host defense responses known to occur on CF airways.

### An Immature Proinflammatory Archetype Prevails Among CF Airway PMN

CF Sputum contained 64% PMN, in contrast with HC where PMN constituted 2% of sputum cells (Fig. 1D). PHATE of the PMN spectrum of gene expression (PMN manifold) enabled us to identify three PMN archetypes based on canonical markers of PMN immaturity (*CXCR4, IGF2R*) and maturity (*FCGR3B, ALPL, and CXCR2*), as well as a heat-shock response archetype (Fig. 3A, B, Fig. S5). To analyze gradual changes within the PMN manifold, we applied trajectory inference and correlated the resulting pseudotime distances with gene expression and regulon activity. When tracing PMN maturation, we observed that expression of calprotectin (*S100A8, S100A9*), *S100A11, CSF3R* and antiapoptotic factor *BL2A1* are gained relatively early, in contrast to classical maturation markers *FCGR3B, ALPL, CXCR2* and *CD14* which ramp up in expression relatively late (Fig. 3B, Data file S2) (*24*). In immature PMN, we observed a gradual increase of transcription factors *TFEC, MITF, STAT3*, and maturation-associated transcription factors *CEBPB*, and *NFIL3*. The CF-predominant immature PMN archetype was further defined by increased expression of PMN-activating chemokine MIP (*CCL3, CCL4*) and downstream transcription factor and adapter molecules *IRAK3* and *TRAF3*. These findings suggest that CF airway PMNs have an overall proinflammatory phenotype, with a large subpopulation of PMNs exhibiting a functional and maturity transcriptional shift, consistent with an immature PMN gene expression profile.

**Fig.3.**
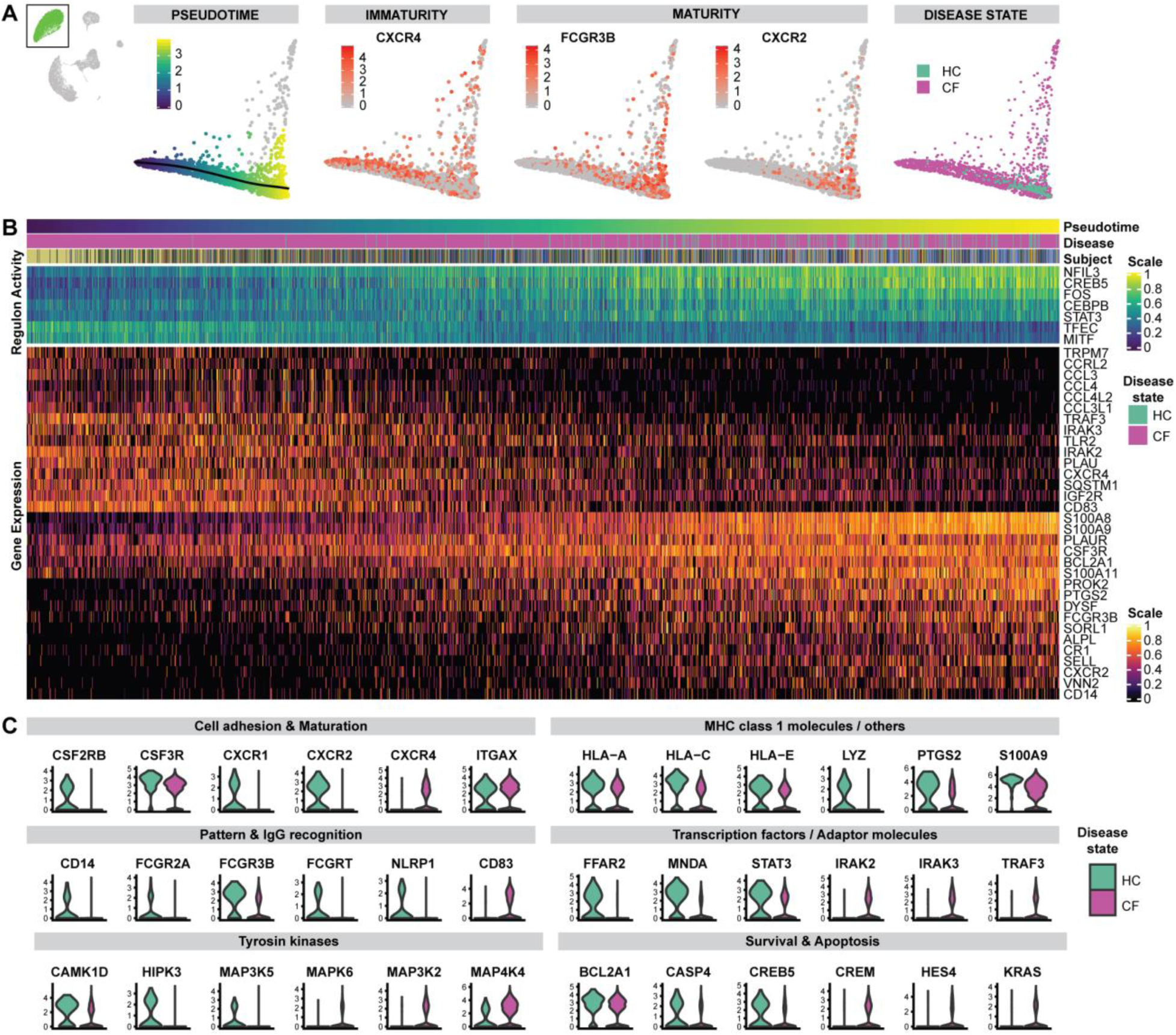
An Immature Proinflammatory Archetype Prevails Among CF Airway PMN. **(A)** PHATEs of PMN, colored by: (i) pseudo time from immature to mature PMNs, (ii) examples of canonical marker features of immaturity (CXCR4) and maturity (FCGR3B, CXCR2) in peripheral PMN, (iii) disease state. The cells deviating upward are PMN expressing heat-shock response genes, for PHATE embedding colored by gene expression of HSPA1A, HSPH1, and DNAJB1, see Fig. S5A). For corresponding PHATE embedding colored by disease state and subjects, see Fig. S5B. **(B)** Heatmap of gene expression and regulon activity in PMNs, ordered by pseudotime distances along PHATE manifolds that transition from CF-enriched regions of immature and activated PMN archetype towards control-enriched mature PMN archetype. Annotation bars represent the pseudotime distance, disease status, and subject for each cell; expression values are centered and scaled. **(C)** Violin plots of differentially expressed genes comparing CF and control PMN populations (for p-values see Data file S3), grouped by disease state, and sorted thematically.

### CF PMN Archetypes Have Decreased Phagocytic Marker and Tyrosine Kinase Expression

We compared the gene expression profiles of CF and HC PMN to understand transcriptomic differences associated with their immune function (Data file S3). We categorized the top gene expression differences between CF and HC accordingly into: 1) Cell adhesion and maturation markers, 2) MHC class I molecules and phagocytosis; 3) Transcription factors and adapter molecules, 4) Cell survival and apoptotic signaling; 5) Tyrosine Kinase expression; and 6) Hypoxic response (Fig. 3C). In CF PMN, cell adhesion and maturation markers were overall lower than in HC (*CSF2RB, CSF3R, CXCR2, ICAM3, PECAM1*), except for *ITGAX*. The decreased expression of these markers in CF reflects a higher prevalence of the immature PMN archetype described above. In addition to decreased CXCR- and CSF-receptor expression, CF PMN also expressed lower *CXCR1, ILR1*, and *IL1B* that could condition further defects in phagocytosis and inflammatory cell recruitment. We identified striking differences in antigen presentation, pathogen recognition, and phagocytosis-associated genes between CF and HC PMN. CF PMN showed decreased expression of numerous members of the MHC-I molecules (*HLA-A/B/C/E*), immunoglobin receptors (*FCGR3B, FCGR2A, FCGRT*), decreased pathogen recognition receptors *CD14, TLR2*, and *NLRP1*, and decreased expression of lysozyme (*LYZ*). Interestingly, two genes involved in the assembly of lipid rafts and primary neutrophil granule release were increased (*SYK, CD63*) suggesting that although PMN may suffer from defective phagocytic activity, the transcriptional infrastructure needed to express tissue proteases and inflammatory mediators into the airways is preserved. CF PMN demonstrated increased transcriptomic activation characterized by expression of transcription factors and proinflammatory adapter molecules (increased *PI3, IRAK2/3, TRAF3, TANK*), yet this activation did not translate into increased expression of inflammatory cytokines. Interestingly, the downstream response to cytokine activation appeared to be blunted, as shown by decreased overall tyrosine kinase gene expression (*ITPK1, MAP3K5, MAP2K4, CAMK1D, PIK3CD, HIPK3*). Finally, we observed the induction of genes involved in the hypoxic response (*HIF1A, VEGFA, FGF13, PTGS2*) and diverging proapoptotic signals with lower expression of *CASP4, RPS6KA5, CREB5, and BCL2A*, and increased expression of *HES4, KRAS*, and *CREM* in CF. These observations underscore the presence of a hypoxic airway environment in CF and a dysfunctional cell death program that enhances the survival of functionally ineffective PMN. Taken together, these findings indicate that CF PMN do not carry out an effective transcriptional response to inflammatory stimuli and lack essential components for pathogen recognition and removal.

## Discussion

This is the first single-cell transcriptome characterization of immune cells in CF sputum. We identified CF-specific differences in cell subpopulations including alvMΦ, RLPs, and PMN. Furthermore, these cells had markedly different transcriptional profiles when compared to their HC counterparts. Previous CF studies have used transcriptomic analysis to determine the likelihood of adverse outcomes in CFLD, however they have not focused on establishing differences between healthy and CF airway inflammatory cells, or characterizing their immune activation profiles (*12*). The most remarkable finding from this study is the discovery of novel archetypes of RLPs, enabled by an unprecedented depth of gene expression profiling. These inflammatory cell subpopulations exhibit a wide spectrum of maturity and immune activation in CF. Airway MΦ and other mononuclear cells have been described in human CF airway secretions (*16, 17*) and their role in driving exaggerated airway inflammation in CF has been well characterized in animal models (*4, 9, 16, 19*). However, a broader genomics approach to define sputum RLPs, their potential functional impairments, and pathogenic role has not been reported.

We identified three archetypes of CF RLP. These archetypes include activated Mo, mature MoMΦ, and HS-Mo. Airway Mo in CF have impaired ion transport and phagocytic function, however their role in CFLD remains undefined (*20, 25*). Others have described dramatic changes in Mo cell adhesion and chemotaxis that perpetuate inflammation in CF lungs, along with enhanced chemokine production that sustains PMN recruitment and injury (*26*). In agreement with these studies, we observe that Mo are rather abundant in CF sputum, but are deficient in Mo maturation gene expression markers (*MITF, SPI1*). Furthermore, CF Mo were not only abundant, but also highly active from the immune perspective, expressing high levels of inflammation-related genes (*CXCL8, IL1B, CCL3*, and Calprotectin). These observations underscore a defect in CF Mo maturation that preserves a highly proinflammatory phenotype and contributes to airway damage and aberrant inflammatory cell recruitment (*18, 27*). In contrast to CF airway Mo, more mature CF phagocytes (MoMΦ, alvMΦ), showed low levels of immune activation markers observed in CF Mo, and of key phagocytic and cytolytic components of the immune response (complement *C1Q, MARCO*). This underscores that in CF, RLPs that reach maturity exhibit transcriptomic evidence of impaired or limited phagocytic function, accounting for the known impaired phagocytic abilities of these cells in CF.

PMN were the most abundant immune cells in the sputum of patients with CF. This is consistent with reports in the CF literature, similar to the predominance of alvMΦ in HC sputum (*14, 15*). However, scRNAseq enabled Pseudotime and regulon analyses that led to the discovery of new archetypes of CF PMN based on inflammatory and maturity gene expression markers. One, characterized by high maturity and a proinflammatory transcriptional state, and another with lower proinflammatory activity and delayed expression of maturity markers. Overall, the increased expression of proinflammatory genes in mature PMN highlights a highly activated and proinflammatory state, clearly distinguishable from the transcriptional profile of HC PMN. The immature airway PMN archetype shares features of a previously described subpopulation of transmigrated PMN with increased granule release, immunoregulatory and metabolic activity, and defective bacterial killing in in vitro studies, referred to as GRIM neutrophils (*10, 11, 28*). We identified cells with similar characteristics, but as part of a spectrum of granulocyte maturation that encompasses vigorously activated PMN on one extreme and PMN with decreased expression of maturity markers & evidence of recent migration into the airways on the other extreme. Adding to the complexity of these PMN subpopulations, counterproductive pro- and anti-apoptotic signals were present across the CF PMN when compared to HC (increased *UVRAG, PLPP3, ATG7*, decreased *CASP4, RPS6KA5, CREB5, BCL2A*). Taken together, these findings underscore an aberrant proinflammatory state in CF PMN, exacerbated by disruption of immunomodulatory and anti-inflammatory mechanisms like apoptosis and transcription factor suppression.

This work includes two technical advances. First, this is the only reported scRNAseq study of CF sputum, a notoriously complex biological sample with high variability in cell viability and in cellularity between subjects. Second, our sputum processing protocol avoids the use of reducing agents to solubilize sputum and instead minimizes immune cell activation and injury by using mechanical disruption and filtering. Importantly, ours is the first report of a sputum cryopreservation protocol allowing the retrieval of live cells for scRNAseq analysis. The ability to use cryopreserved cells overcomes a major limitation of previous single-cell studies that required fresh samples (*22, 29*), this is particularly important for the recovery of PMN, known for their short life-span ex-vivo and susceptibility to immune activation (*30*). Our study has several limitations: 1) Large differences in predominant cell types between CF and HC subjects make it difficult to generalize gene expression changes between disease and control groups. Although we present these comparisons, our focus is on understanding CF-specific cell distributions and their spectrum of maturity and activation markers; 2) Since HC express minimal sputum if any at all, we used a standardized approach for sputum induction in these subjects, while CF cells were obtained from spontaneously expectorated sputum. As single cell suspensions are standardized for number of cells before any analysis, these sampling differences likely have a minor impact on our observations; finally, 3) Our study has a small sample size; however, we sought to match subjects according to age and sex, and HC were compared to a relatively homogeneous CF cohort in terms of *CFTR* mutation background, CF comorbidities, and ongoing therapy. Although a small number of patients were recruited for this study, we believe they are representative of patients with CF based on the *F508del* allele frequency in our cohort and the identification of nine distinct cell types representative of airway cells in CF. Despite these limitations, our findings are robust and representative of the CF airway compartment.

CF research has rapidly progressed towards clinical, molecular, and functional characterization based on individualized high-throughput diagnosis and functional profiling. Our application of scRNAseq enabled the discovery of transcriptional archetypes in CF-specific cell subpopulations that may underlie subject-specific differences in disease progression and response to therapy. As we progress towards increasingly early applications of therapeutic and genomic technologies in CF, we hope that this approach to individualized profiling of airway inflammation may serve as a foundation for further discoveries that transform the natural history of CFLD.

## Materials and Methods

### Subject Cohort

A total of nine subjects with a confirmed diagnosis of CF from the Yale Adult CF Program provided sputum samples for this study, five during exacerbation and five during periods of stability. These subjects were recruited during a) Scheduled routine visits (n=5) and b) Unscheduled “sick” visits, in which they reported new respiratory symptoms and were diagnosed with a CF exacerbation (n=4). A CF exacerbation was defined by the emergence of four of twelve signs or respiratory symptoms, prompting a change in therapy and initiation of antimicrobial treatment (modified from Fuchs’ criteria (*31*)). These criteria included: change in sputum; change in hemoptysis; increased cough; increased dyspnea; malaise, fatigue or lethargy; fever; hyporexia or weight loss; sinus congestion; change in sinus discharge; change in chest physical exam; or FEV_1_ decrease >10% from a previous value (*31*). Individuals without new symptoms and those that did not meet AE criteria were characterized as “CF Stable”. Our recruitment period extended through 2019. We also recruited five healthy volunteers (Healthy Controls, HC) to undergo sputum induction according to previous protocols (*32*). Since we did not identify significant differences in the gene expression profiles of stable and exacerbation subjects, all CF subjects were grouped as “CF” as compared to healthy control samples for analysis as a group. The study protocol was approved by the Yale University Institutional Review Board and informed consent was obtained from each subject.

### Sputum Collection and Processing

CF subjects expectorated sputum spontaneously for our studies. Induced sputum samples were obtained from HC as previously described (*32, 33*). Briefly, subjects inhaled nebulized 3% hypertonic saline for five minutes on three cycles. To reduce squamous cell contamination, subjects were asked to rinse their mouth with water and clear their throat. Expectorated sputum samples were collected into specimen cups and placed on ice. Sputum plug material from HC and CF subjects were selected and weighed prior to washing with 9x their volume of PBS. Samples were incubated in Dulbecco’s Phosphate-Buffered Saline (DPBS) with agitation for 15 minutes and filtered through 40-micron strainers. Samples were centrifugated at 300 g for five minutes and supernatants were stored at -80°C. The pellets were suspended in RPMI/10%FBS medium with 10% DMSO. Aliquots of 1 ml was saved into cryogenic vials and placed in Nalgene Cryo 1° C Freezing Container (Sigma, St. Louis, MO) overnight at -80°C. Samples were stored in liquid nitrogen the next day. Frozen samples were thawed in a water bath at 37°C, resuspended with 20ml DMEM + 10% heat-inactivated FBS (Life Technologies, USA), then centrifuged at 300g, 5min, 4°C. Supernatant was discarded, cells were resuspended in 2ml DMEM + 10% FCS, passed through a 70µm cell strainer (Fisher Scientific, USA). Non-viable cells and mucus were removed from the cell suspensions using a OptiPrep (Iodixanol) density gradient centrifugation according to the manufacturer’s protocol (OptiPrep Application Sheet C13 – Strategy 2). In brief, 1·86ml of the cell suspensions were mixed with 40% OptiPrep in DMEM + 10% FCS by repeated gentle inversion, overlaid with a density barrier (density: 1·09g/ml, 780µl OptiPrep in 2.22ml DMEM + 10% FCS), then overlaid with 500µl DMEM + 10% FCS. After centrifugation at 800g, 20min, 4°C, viable cells were collected from the top interface and diluted with 2ml DMEM + 10% FCS, centrifuged at 400g, 5min, 4°C, then resuspended in 1ml PBS + 0.04% BSA (New England Biolabs, USA) and passed through a final 40µm cell strainer (Fisher Scientific, USA). For cell concentrations, cells were stained with Trypan blue and counted on a Countess Automated Cell Counter (Thermo Fisher, USA).

### Single Cell Barcoding, Library Preparation, and Sequencing

Single cells were barcoded using the 10x Chromium Single Cell platform, and cDNA libraries were prepared according to the manufacturer’s protocol (Single Cell 3’ Reagent Kits v3, 10x Genomics, USA). In brief, cell suspensions, reverse transcription master mix and partitioning oil were loaded on a single cell “B” chip, then run on the Chromium Controller. mRNA was reverse transcribed within the droplets at 53°C for 45min. cDNA was amplified for a 12 cycles total on a BioRad C1000 Touch thermocycler. cDNA was size-selected using SpriSelect beads (Beckman Coulter, USA) with a ratio of SpriSelect reagent volume to sample volume of 0.6. For qualitative control purposes, cDNA was analyzed on an Agilent Bioanalyzer High Sensitivity DNA chip. cDNA was fragmented using the proprietary fragmentation enzyme blend for 5min at 32°C, followed by end repair and A-tailing at 65°C for 30min. cDNA were double-sided size selected using SpriSelect beads. Sequencing adaptors were ligated to the cDNA at 20°C for 15min. cDNA was amplified using a sample-specific index oligo as primer, followed by another round of double-sided size selection using SpriSelect beads. For qualitative control purposes, final libraries were analyzed on an Agilent Bioanalyzer High Sensitivity DNA chip. cDNA libraries were sequenced on a HiSeq 4000 Illumina platform aiming for 150 million reads per library. Full de-identified sequencing data for all subjects is available in the gene expression omnibus (GEO) under accession number GSE145360.

### Data Processing and Computational Analyses

Basecalls were converted to reads with the implementation mkfastq in the software Cell Ranger (v3.0.2). Read2 files were subject to two passes of contaminant trimming with cutadapt (v2.7): first for the template switch oligo sequence (AAGCAGTGGTATCAACGCAGAGTACATGGG) anchored on the 5’ end; secondly for poly(A) sequences on the 3’ end. Following trimming, read pairs were removed if the read 2 was trimmed below 20bp. Subsequent read processing was conducted with the STAR (v2.7.3a) (*34*) and it’s single cell sequencing implementation STARsolo. Reads were aligned with to the human genome reference GRCh38 release 31 (GRCh38.p12) from GENECODE (*35*). Collapsed unique molecular identifiers (UMIs) with reads that span both exonic and intronic sequences were retained as both separate and combined gene expression assays. Cell barcodes representative of quality cells were delineated from barcodes of apoptotic cells or background RNA based on the following three thresholds: at least 10% of transcripts arising from intron spanning, i.e. unspliced reads indicative of nascent mRNA; more than 750 transcripts profiled; less than 15% of their transcriptome was of mitochondrial origin. Technical summaries related to sequencing and data processing can be found in Data file S4.

### Data Normalization and Cell Population Identification

UMIs from each cell barcode - irrespective of intron or exon coverage - were retained for all downstream analysis and analyzed using the R package Seurat (version 3.1.1) (*36*). Raw UMI counts were normalized with a scale factor of 10,000 UMIs per cell and subsequently natural log transformed with a pseudocount of 1. More than double the cell barcodes were detected in two subjects compared to all other subjects, so cells were randomly downsampled to a maximum of 2,250 cells per subject to avoid predominance of those two subjects. 3000 highly variable genes were identified using the method “vst”, then data was scaled and the total number of UMI and the percentage of UMI arising from mitochondrial genes were regressed out. The scaled values were then subject to principle component analysis (PCA) for linear dimension reduction. A shared nearest neighbor network was created based on Euclidean distances between cells in multidimensional PC space (the first 12 PC were used) and a fixed number of neighbors per cell, which was used to generate a 2-dimensional Uniform Manifold Approximation and Projection UMAP for visualization. For cell type identification, scaled data was clustered using the Leiden algorithm. In addition to general filtering based on quality control variables, a curated multiplet removal based on prior literature knowledge was performed: Cell barcodes were identified as mulitplets if their expression level was higher than 1 in the following marker genes (outside the appropriate cluster): MS4A1 (B cells), CD2 (T cells), VCAN (monocytes), FCGR3B (neutrophil granulocytes), KRT19 (epithelial), and FABP4 (alveolar macrophages). Cell barcodes flagged as multiplets were not included in downstream analyses.

### Generation of Cell Type Markers and Differential Expression Between Disease Conditions

In order to evaluated cell-type markers we used Seurat’s FindAllMarkers (to calculate log fold changes, percentages of expression within and outside a group, and p-values of Wilcoxon-Rank Sum test comparing a group to all cells outside that specific group including adjustment for multiple testing) and additionally calculated binary classifier system based on diagnostic odd’s ratios as described in our earlier work (*23*) (Data file S2). For each cell type in the data, we identified the genes whose expression was log fold change >= 0·25 greater than the other cells in the data. We then calculated the diagnostics odds ratio (DOR) for each of these genes, where we binarize the expression values by treating any detection of a gene (normalized expression value > 0) as a positive value, and zero expression detection as negative. We included a pseudocount of 0.5 to avoid undefined values, as:

DOR = ((TruePositives + 0.5) / (FalsePositives + 0.5)) / ((FalseNegatives + 0.5) / (TrueNegatives + 0.5))

where True Positives represents the number of cells within the group detected expressing the gene (value > 0), FalsePositives represents the number of cells outside of the group detected expressing the gene, FalseNegatives represents the number of cells within the group with no detected expression, and TrueNegatives represents the number of cells outside of the group with no detected expression of the gene. For differential expression testing between disease conditions, Seurat’s implementation of a Wilcoxon-Rank Sum in FindMarkers was used, only testing genes whose expression was log fold change >= 0·25 greater between both disease conditions.

### Scoring of regulon activity and pathways

A regulon is defined as a group of target genes regulated by a common transcription factor. To score the activity of each regulon in each cell, we utilized the package pySCENIC (*37*) with default settings and the following database: cisTarget databases (hg38 refseq-r80 500bp_up_and_100bp_down_tss.mc9nr.feather, hg38 refseq-r80 10kb_up_and_down_tss.mc9nr.feather) and the transcription factor motif annotation database (motifs-v9-nr.hgnc-m0.001-o0.0.tbl) which were both downloaded from resources.aertslab.org/cistarget/, and the list of human transcription factors (hs_hgnc_tfs.txt) which was downloaded from github.com/aertslab/pySCENIC/tree/master/resources.

In order to calculate pathway activity scores, Gene Ontology (GO; geneontology.org) pathways related to monocyte/macrophage functions were downloaded, then scored using Seurat’s AddModuleScore using default settings.

### Pseudotime Analysis of PMN and monocytes/macrophages

We observed already in UMAP space that many features in the data were represented by a continuum of increasing phenotypic deviation, e.g. increase of maturation markers in neutrophil granulocyte, maturation from monocytes to macrophages, and gradual increase of classical markers of inflammation in monocytes. Consequently, we sought to implement pseudotime analysis of these continua to assess features rather than relying on traditional group-wise comparisons. Cell barcodes were subsetted to either only neutrophil granulocytes or monocytes/macrophages. Due to major differences in number of cells profiled per subject, PMN were randomly downsampled to a maximum of 200 cell barcodes per subject, and in the Mo/MΦ subgroup to a maximum of 250 cell barcodes per subject. As for the full dataset, data of the subgroups was normalized, variable features were extracted (200 for PMN, 500 for Mo/MΦ), scaled, then subject to PC analysis. PHATE (Potential of Heat-diffusion for Affinity-based Trajectory Embedding)(*38*) embedding was performed which is specifically suitable to continua (50 nearest neighbors, 5 PCs, t=50 in Mo/MΦ and t=100 in PMN). Cell barcodes were clustered using the cluster_phate function (k=8) for PMN and the Leiden clustering for Mo/MΦ. Trajectories were identified using Slingshot (*39*) on the PHATE embeddings with default settings, and a central starting cluster for the Mo/MΦ. Pseudotime analysis was used to distinguish gene expression trajectories, and in turn, the most extreme phenotypes of these trajectories defined transcriptional archetypes in sputum (*21, 40, 41*). Pearson’s correlation coefficients and their p values, including Bonferroni adjustment for multiple testing, were calculated between the resulting pseudotime distances of these trajectories and gene expression and the regulon activity scores (Data file S2). Gene expression and regulon activity scores correlating with pseudotime values were visualized by heatmaps.

### Validation of major cell types by Cytometry Time of Flight (CyTOF)

CyTOF-derived fcs files from the study by Yao et al. (*22*) were processed using the bead-based Normalizer Release R2013a (*42*). Normalized files were then processed in Cytobank (https://premium.cytobank.org/) using gates to select singlets, remove beads and identify live cells. Events identified using this workflow were exported and processed further using the R package cytofkit version 1.12.0 (*43*). The Rphenograph function in cytofkit was implemented to cluster cells using cytofAsinh method, with the tsne dimensionality reduction method applied on 80000 events, using k=40. Files were merged using the fixed method and the HLA-DR, CD11b, CD8a, CD20, CD16, MIP-1b, TNFa, CD45, CD4, IL-6, CD11c, CD14, Cytokeratin, CD80, CD15, CD163, IFNg, EGFR, CD66b, IL-8, CD62L and CD56 markers were used in this model. Resulting clusters were manually curated and merged after review of surface marker profiles.

### Correlation matrix of immune cell populations comparing sputum and lung cell populations

To identify classifier genes, differential gene expression of immune cell types of this study and analogue cell types from an independent scRNAseq, a dataset of 28 healthy distal lung samples (*23*) was established using Seurat’s FindAllMarkers with an absolute log fold change threshold of 1 (the lung dataset was downsampled within the FindAllMarkers function using the settings: max.cells.per.ident=1000, seed=7). Classifier genes were filtered such that all genes had a Bonferroni adjusted p-value < 1E-5. For each cell type and each dataset, the top 50 marker genes, ordered by fold change, were selected. We took the intersection of the genes from both datasets as top classifiers (n=154). The average gene expression of these 154 genes were calculated for each cell type per dataset. Spearman correlation matrix was calculated using base R’s function “cor”. The R package “corrplot” was used to visualize the Spearman correlation matrix. Unsupervised hierarchical complete clustering was performed to order the cell types in the heatmap.

## Data Availability

Sequencing data was deposited on the gene expression omnibus (GEO) under accession number GSE145360.

https://www.ncbi.nlm.nih.gov/geo/query/acc.cgi?acc=GSE145360

## Acknowledgments

We thank our patients, the medical staff at the Yale Adult Cystic Fibrosis Program, Dr. Farida Ahangari, and Dr. Jonathan Koff, Director of the Yale Adult CF Program, for their support and contributions to this project. Sequencing was conducted by Mei Zhong at Yale Stem Cell Center Genomics Core facility which was supported by the Connecticut Regenerative Medicine Research Fund and the Li Ka Shing Foundation.

## Author contributions

CJB and NK conceptualized, acquired funding and supervised the study. CJB, SK, and MEE performed sample collection, phenotyping, and sputum processing. GLC facilitated sputum collection infrastructure and processing protocols. JCS and TSA performed single cell barcoding library construction. Data was processed, curated and visualized by JCS under the supervision of XY, CJB, and NK, and analyzed by JCS, EMB, MS, CSD, and CJB. CyTOF data were reanalyzed by JLG, RRM, and EMB. JCS, TSA, PS, IOR, and NK created and provided scRNAseq data of control distal lungs, TSA calculated the correlation matrix. The manuscript was drafted by JCS and CJB, and was reviewed and edited by all other authors.

## Supplementary Materials

**Fig. S1.**
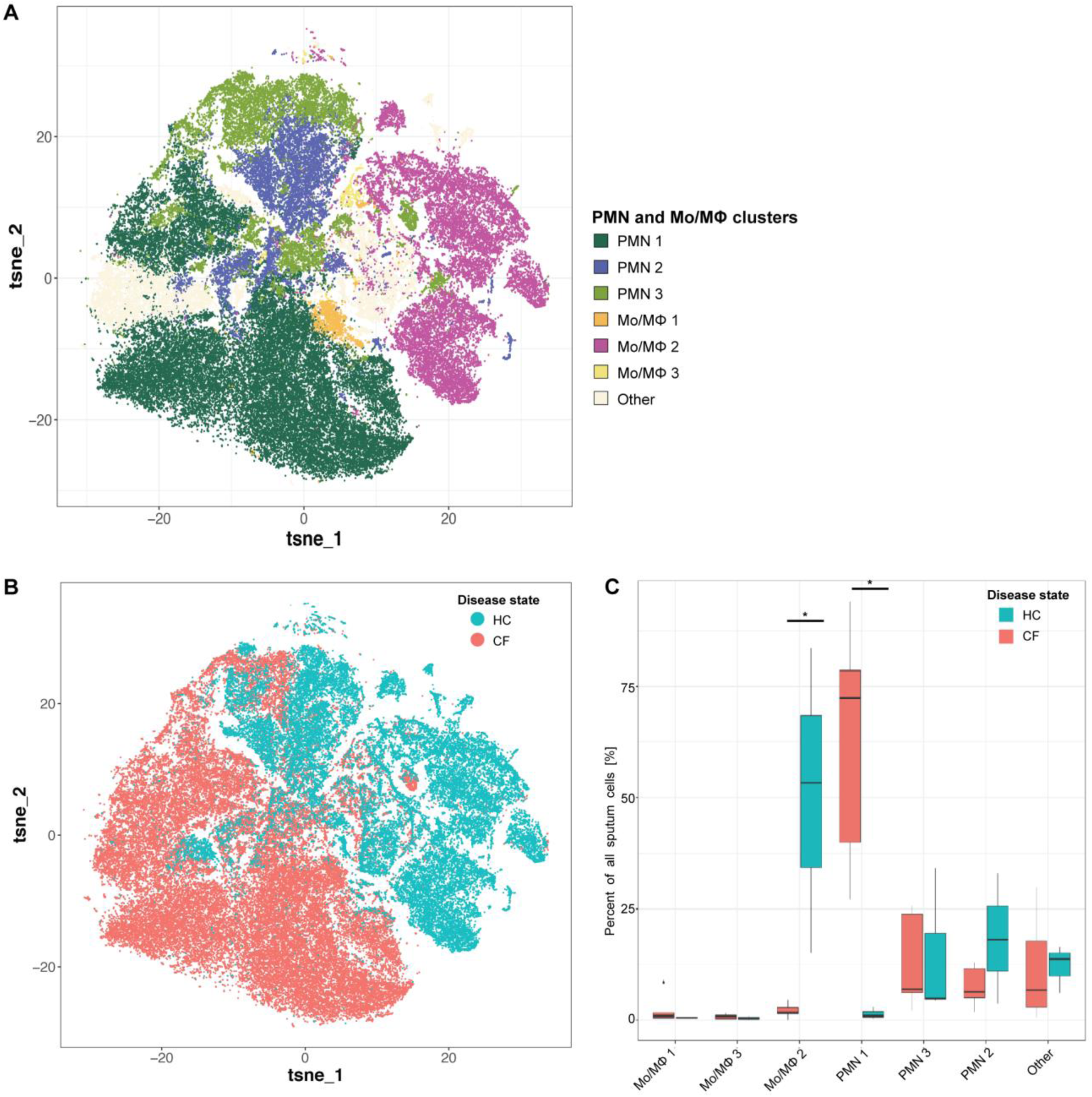
Validation of the shift of major immune cell types in sputum of CF compared to HC. **(A)** RPhenograph clustering of Sputum CyTOF in patients with cystic fibrosis (CF) and healthy controls (HC) demonstrates differences in the populations of immune cells. The sputum of patients with CF is characterized by high percentages of neutrophils, while sputum from HC is characterized by high percentages of macrophages. **(B)** RPhenograph clustering of Sputum CyTOF according to Healthy Control (HC) and Cystic Fibrosis (CF) status. **(C)** Boxplots showing percentages of Mo/MΦ, PMN, and other to all cells profiled per subject, separated by disease state. Whiskers represent 1.5 × interquartile range (IQR). * p < 0.05 determined by a Wilcoxon rank sum test comparing cell percentages of CF patients and controls.

**Fig. S2.**
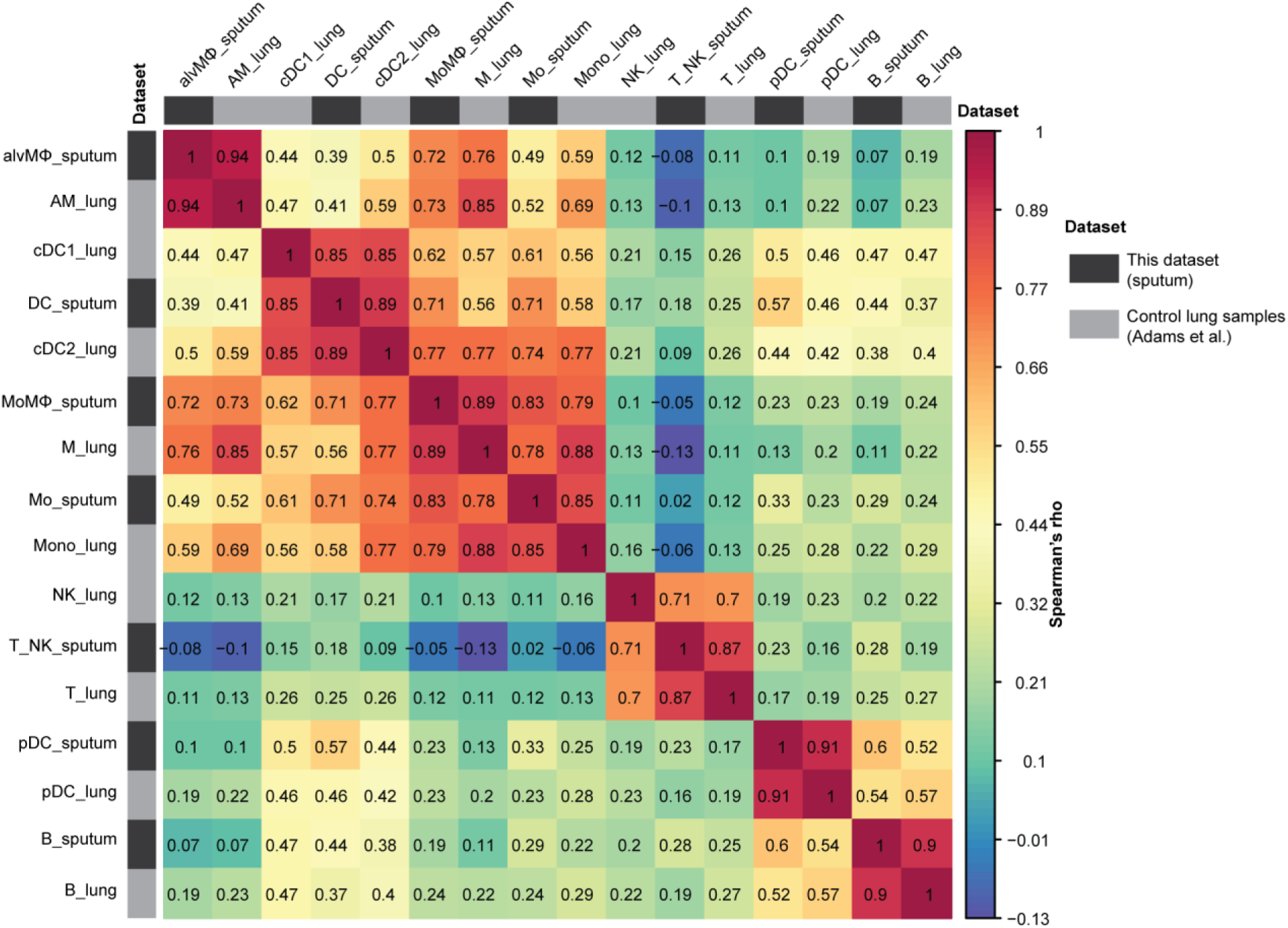
Concordance of cell type annotations. Correlation matrix of immune cell populations of this study and analogous cell types from an independent scRNA sequencing dataset of distal lung samples, subsetting to the 28 healthy controls. Matrix fields are colored by Spearman’s rho, cell types are ordered by unsupervised hierarchical clustering. Annotation bars are highlighting the two different datasets (dark grey: this dataset, light grey: lung samples from healthy controls only from Adams, et al. (*23*)).

**Fig. S3.**
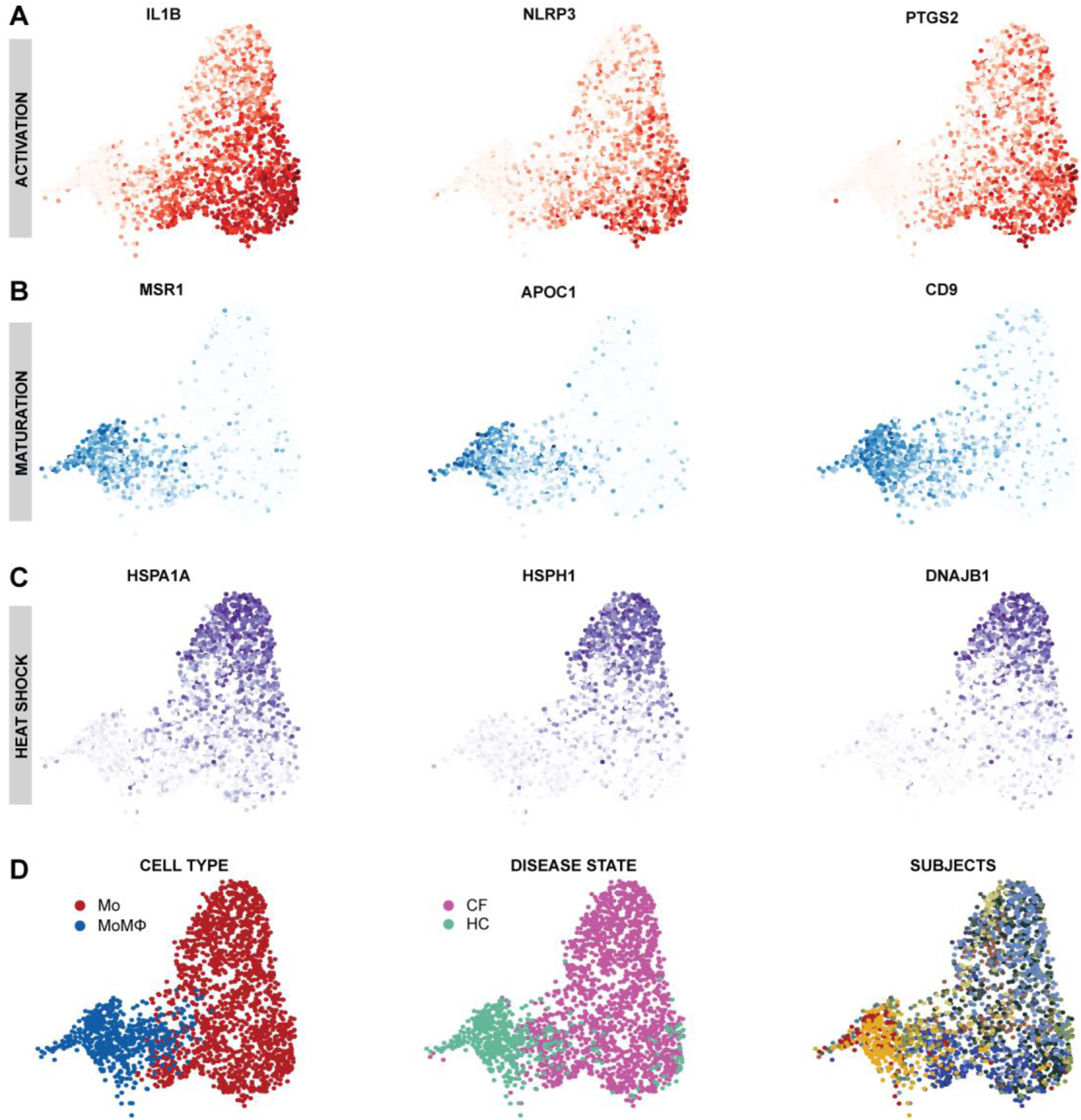
Expression of selected marker genes of Mo/MoMΦ trajectories on UMAPs. **(A)** UMAP, zoomed in on Mo and MoMΦ, colored by expression of inflammatory genes IL1B, NLRP3, PTGS2. **(B)** UMAP, zoomed in on Mo and MoMΦ, colored by expression of mature macrophage genes MSR1, APOC1, CD9. **(C)** UMAP, zoomed in on of Mo and MoMΦ, colored by expression of heat shock genes HSPA1A, HSPH1, DNAJB1. **(D)** UMAP, zoomed in on of Mo and MoMΦ, colored by (i) cell type, (ii) disease state, (iii) subjects. CF: Cystic Fibrosis, HC: Healthy Control, Mo: Monocyte; MoMΦ: monocyte-derived macrophage.

**Fig. S4.**
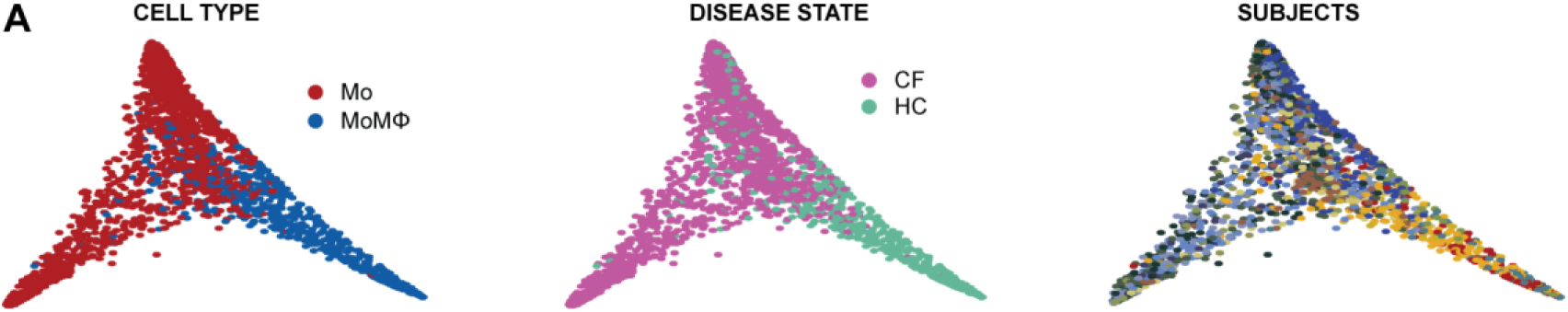
Additional annotations of Mo/MoMΦ on PHATE embedding. **(A)** UMAP of Mo and MoMΦ colored by (i) Cell type, (ii) Disease state, (iii) Subjects. CF: Cystic Fibrosis, HC: Healthy Control, Mo: Monocyte; MoMΦ: monocyte-derived macrophage

**Fig. S5.**
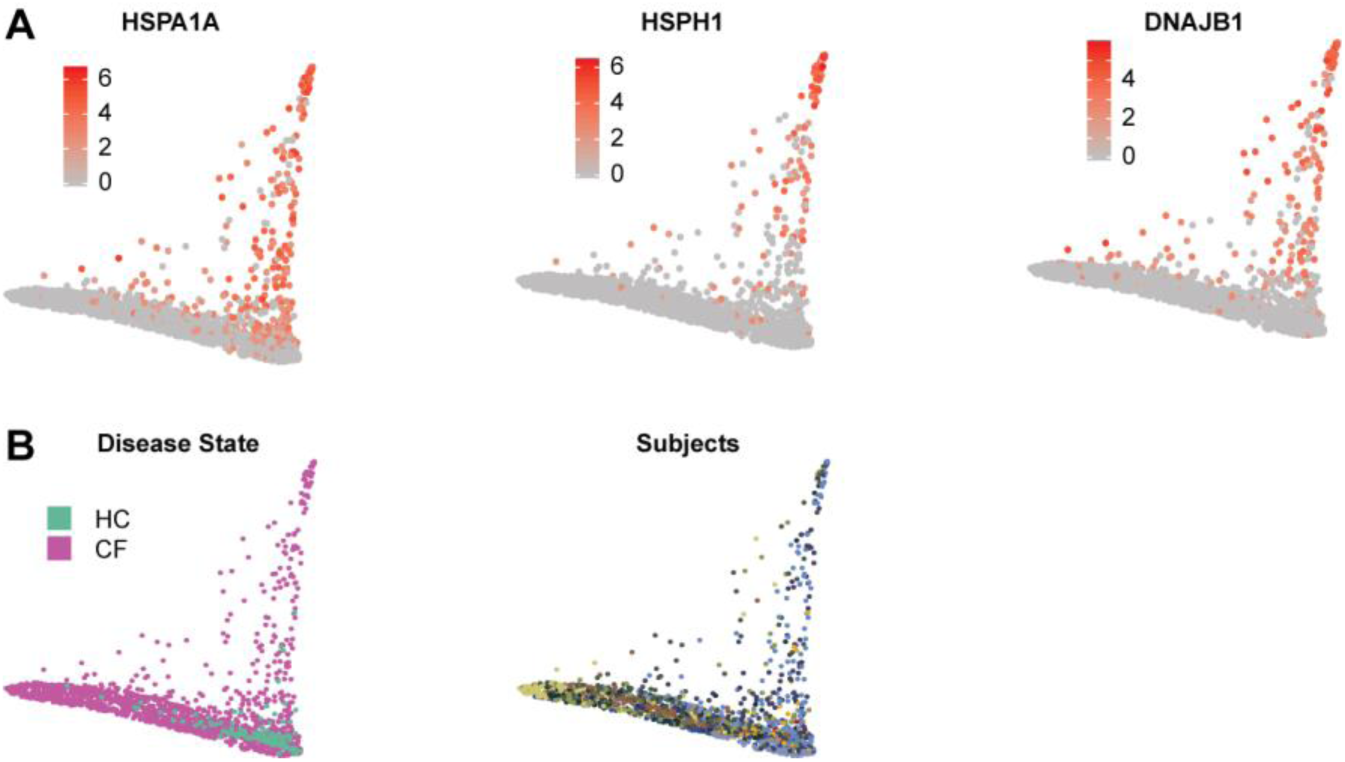
Additional annotations of PMN on PHATE embedding. **(A)** PHATE of PMN colored by expression of heat shock genes HSPA1A, HSPH1 and DNAJB1. **(B)** PHATE of PMN colored by disease state (HC: Healthy Control, CF: Cystic Fibrosis) and subjects.

**Data file S1**. Results of Wilcoxon rank-sum test and log transformed diagnostics odds ratio of genes for cell types, subsetting to genes with log transformed fold change > 0.25 for each cell population compared to all other cell populations.

**Data file S2**. Results of Pearson correlation between gene expression and pseudotime distance values within each trajectory.

**Data file S3**. Results of Wilcoxon rank-sum test on gene expression within each cell type comparing CF to HC.

**Data file S4**. Technical summary of all sequenced and processed libraries of this dataset. TSO: template switch oligo.

